# Evolutionary rescue effect disappears under more realistic assumptions

**DOI:** 10.1101/2023.05.19.23290032

**Authors:** Martin Hinsch, David L. Robertson, Eric Silverman

## Abstract

In a recent paper Zhang et al. [1] elegantly incorporate the evolution of inter-host virus fitness into an epidemiological model. They show that this leads to substantial changes in the system dynamics and in particular that evolution can “rescue” the pathogen population if the mutation rate is high enough. However, their model rests on the assumption that mutations that affect inter-host fitness are neutral on average. Here we show that under more realistic assumptions concerning the fitness distribution of mutations, the effect can easily disappear and higher mutation rates in fact reduce the likelihood of a pandemic.

## 1 Introduction

Zhang et al. [1] present an analytical epidemiological model, backed by numerical simulations, that takes into account the evolution of intra- and inter-host virus fitness. Their results show that in situations with *R*_0_ *<* 1 that would classically be assumed to lead to the eventual disappearance of the pathogen, under high mutation rates evolution can increase *R*_0_ quickly enough to prevent extinction. Their model rests on the assumption that mutations affecting interhost fitness are neutral for intra-host fitness (transmission) and equally likely to have positive or negative consequences for inter-host fitness. If this is the case, their accumulated effect over the time span between first infection and transmission can therefore be modelled as a random walk, leading at point of transmission to a normal distribution of inter-host fitness with the mean at 1 and the standard deviation representing mutation rate.

As a general rule, however, adaptive mutations are usually rare compared to those with no or negative effects on fitness [2], a pattern that seems to hold in virus populations as well [3]. This was exemplified in the COVID-19 pandemic, where the subset of mutations that have emerged giving rise to more transmissible variants, e.g., Spike D614G and the sets of mutations comprising variants of concern (VOCs) were much more significant than recurring mutations both for intra- and inter-host fitness [4]. With that in mind, we replicated the study’s results using a more mechanistic model of the pathogen’s fitness that allows us to adjust the distribution of the effect of mutations on inter-host fitness.

We are able to replicate the results of Zhang et al. when we incorporate the same assumption of net neutral fitness effects (Figure 1A). However, when we assume more realistically that the effect of mutations is more often negative than positive, pandemic behaviour arises only for high *R*_0_ and low mutation rate. In particular, the evolutionary “rescue effect” that occurs in the original scenario, disappears nearly entirely and higher mutation rates prevent rather than trigger pandemic behaviour (Figure 1B). We note that the distribution of fitness effects we assume in this case is much closer to neutrality than the distributions found empirically [3].

**Figure 1:**
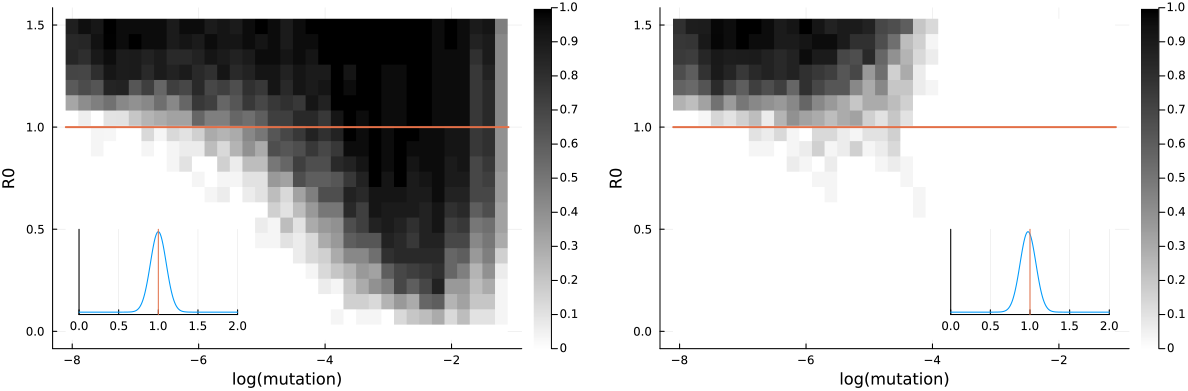
Phase diagram for unbiased (*μ* = 1.0, left) and slightly biased (*μ* = 0.975, right) mutation effects. Shading represents mean incidence over 30 replicates after 3000 time steps.

Zhang et al. provide a simple and elegant illustration of a mechanism by which a sub-pandemic pathogen may ‘break through’ into a pandemic phase driven by mutation. Our replication demonstrates that a mechanistic approach can reproduce the results of the original model, but also that these results might only apply under specific assumptions. The scenarios presented in the supplementary material of the original paper suggest a similar conclusion. Here, assuming a correlation between inter- and intra-host fitness leads to similar large-scale changes in the system behaviour (as shown by the phase diagrams in Figure S4). However, contrary to our results, in this case higher mutation rates increase rather than reduce the probability of pandemic behaviour.

It seems therefore that while the assumption of pathogen evolution can indeed strongly affect under which circumstances a pandemic can occur, how exactly this effect plays out is highly sensitive to initial assumptions regarding the fitness effects of mutations. Future work could build on these foundational results by incorporating the evolution of both intra- and inter-host fitness based on realistic assumptions relevant to specific pathogens. We propose that such models could provide more insightful long-term forecasting of the epidemiological dynamics of evolving pathogens.

## Methods

We implemented an agent-based model in Julia replicating the original agent-based version of the model (see section Methods in the original paper) where possible, that is in particular the assumptions concerning number of agents, network topology and initial number of infected agents.

In their implementation Zhang et al. model the effects of neutral genetic drift on inter-host fitness as an unbiased random walk by adding a number drawn from a normal distribution to a single fitness value per agent in each time step. We replaced this part of the model with a simple mechanistic model of the occurrence of mutations and their effect on fitness.

We assume that there are only point mutations, that they happen with a fixed probability per base pair and time step and that the same base pair never mutates twice. We can then approximate the number of mutations occurring in a given time step by drawing from a Poisson distribution with parameter *λ* = *p*_mut_ · *n*_bpair_. With the fitness effect of a single mutation *ψ*_*i*_ ∼ *𝒩*(*μ, σ*) the overall fitness after a number of time steps is then simply the product of the initial fitness and the fitness effects of all mutations that have occurred since then:

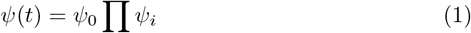

By simulating a large “population” of virus particles over a sufficiently long time span in the way described above we can numerically approximate the probability density function of inter-host fitness at each time step. We then use the expected value of inter-host fitness for a given host - determined by the host’s time since infection *t* and the fitness of the original, infecting pathogen - to calculate that host’s infection probability:

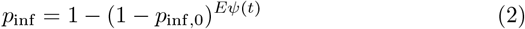

When determining the fitness value for the newly infected host 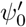 we assume that the distribution of *ψ*(*t*) (equation 1) describes the infecting host’s virus population and that the probability for a given virus particle to cause an infection is proportional to its fitness:

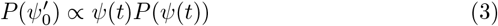

## Data Availability

All code and data produced in the present study are available online and linked to in the manuscript.

https://github.com/mhinsch/virusevolution_NC

## Data availability

The source code repository (see below) contains the model output used to generate the figures.

## Code availability

The source code for our version of the model and all analysis scripts are available at https://github.com/mhinsch/virusevolution_NC.

## Acknowledgements

David Robertson acknowledges funding from the Medical Research Council (MRC, MC UU 12014/12). Martin Hinsch and Eric Silverman are part of the Complexity in Health Improvement Programme supported by the Medical Research Council (MC UU 00022/1) and the Chief Scientist Office (SPHSU16).

This work was also supported by UK Prevention Research Partnership MR/S037594/1, which is funded by the the UK Research Councils, leading health charities, devolved administrations and the Department of Health and Social Care.

We would also like to thank Xiyun Zhang for helpful comments on an earlier version of the manuscript.

## Contributions

M.H. performed the coding and analysis and wrote the manuscript, all with input from D.R. and E.S.

